# Participant Flow Diagrams for Health Equity in AI Research

**DOI:** 10.1101/2023.10.05.23296611

**Authors:** Jacob G. Ellen, João Matos, Martin Viola, Jack Gallifant, Justin Quion, Leo Anthony Celi, Nebal S. Abu Hussein

## Abstract

Biases in sample creation can arise at any study phase, including initial patient recruitment, exclusion criteria, input-level exclusion and outcome-level exclusion, and often reflect the underrepresentation or exclusion of demographic groups historically disadvantaged in medical research. The use of non-representative samples to construct clinical algorithms in artificial intelligence (AI) and machine learning (ML) applications may further amplify this selection bias. Building on the “Data Cards” initiative for transparency in AI research, we advocate for the addition of a detailed participant flow diagram for AI studies, emphasizing the need to detail excluded participant demographic characteristics at every study phase. This tracking of excluded participants enhances understanding of potential algorithmic biases before their clinical implementation, and thus deserves to be detailed in any medical AI study. We include both a model for this flow diagram as well as a brief case study explaining how it could be implemented in practice. Through standardized reporting of participant flow diagrams, we can better gauge the potential inequity embedded in AI applications, facilitating more reliable and equitable clinical algorithms.

## Introduction

An appropriate patient sample is essential to the integrity of any type of medical research. In observational studies, using an inappropriate sample can result in spurious associations unable to be replicated in prospective work. In randomized controlled trials (RCTs), testing an intervention on a non-representative group of patients can result in gaps between the efficacy of a treatment as observed in a trial and its effectiveness in clinical practice. Yet sample biases, often rooted in the historical exclusion of certain demographic groups, including older adults [1], patients with low socioeconomic status (SES) [2], racial minorities and women [3], tend to be consistent and pervasive. These exclusions are reflective of broader structural disparities in the American healthcare system, which have been highlighted by the Joint Commission as targets for assessment of quality of care [4] and by the Centers for Medicare and Medicaid Services as factors in determining reimbursement [5].

Each step of the sample selection process, from initial patient recruitment to exclusion criteria and patient attrition, holds both promise and peril for creating a representative sample. While strategies to create more inclusive, diverse and equitable clinical trial recruitment and designs have been developed in response to these failures [6]–[9], less work has focused on how samples can be fundamentally changed by the steps that follow recruitment, and how these changes can propagate both structural and statistical bias.

As clinical artificial intelligence (AI) and machine learning (ML) applications surge in use, there is the danger that these tools will inherit these biases if trained on nonrepresentative datasets or if they fail to address these potential pitfalls of sample development. Many, for example, have highlighted instances of algorithmic discrimination [10]–[12], which often results from a lack of population representativeness or from the algorithm encoding structural biases already present in a dataset. An additional and under-explored dimension in AI studies is the input-level exclusion of patients due to poor-quality or missing data, which can lead to bias if there is a non-random disparity in data quality or availability among groups. Yet despite the potential for selection bias to propagate within AI algorithms, there are no standardized protocols for reporting participant characteristics and sample creation in medical AI research.

The introduction of “Data Cards” is a recent initiative aimed at changing this and enhancing transparency in AI studies by providing standardized details about a dataset’s background, origin, and purpose [11, 12]. These cards offer an in-depth overview of a dataset, encapsulating 31 distinct facets, which include a variable list, descriptive statistics, and information about its intended application. Building on prior endeavors to boost AI transparency [15]–[18], Data Cards are meant for general use in any AI project and are not tailored specifically for medicine. Thus, while they provide highly valuable insights for medical datasets, their approach could be enhanced by an additional component that meticulously tracks the evolution of patient cohort composition throughout all phases of study sample selection. Many medical studies employ tools like the CONSORT diagram for RCTs [19] and the STROBE diagram for observational studies [20] to accomplish this type of tracking. While these tools are useful, they primarily capture shifts in cohort size and do not track shifts in sample composition. Moreover, a dedicated tool that suits AI studies specifically is needed.

In this article, we advocate for the integration of a detailed participant flow diagram into the current Data Card framework for AI-based medical studies, enhancing its pursuit of transparency and promotion of health equity. In this flow diagram, we argue that it is essential not only to track the number of participants excluded at each phase of any study, but also to report changes in sociodemographic and clinical characteristics relevant to the study question. By doing so, we aim to mitigate and better understand potential statistical and structural biases in the application of AI. To illustrate the importance of tracking excluded participant characteristics through a flow diagram, we walk through various examples that demonstrate how biases can present in different stages of a study, from recruitment to exclusion criteria to input-level omissions to participant attrition during a study. Finally, we present a model for this updated flow diagram and an example of its implementation, envisioning this style of diagram not only as an augmentation to the Data Card but also as an element in medical AI studies generally. While we focus on the use of this flow diagram for AI-based studies here, the concept of tracking cohort composition itself should be encouraged for any type of clinical study.

## Statement of Significance

### Problem

Selection biases generated through the process of cohort composition are often hidden sources of bias that can be propagated by clinical algorithms.

### What is Already Known

Previous studies have identified how sample biases can be generated, and others have detailed their effects on medical AI algorithms. AI models built on non-representative data have been shown to perform poorly for excluded groups, often with implications for health equity.

### What This Paper Adds

This study provides a novel method of tracking how sample composition can change as a study progresses via a standardized participant flow diagram with the aim of ensuring equity and generalizability of results.

## Phases of Sample Selection and Evolution

### Recruitment

While a recruitment population itself cannot be fully captured in a participant flow diagram due to its upstream nature, it is worth addressing how narrow study recruitment can lead to biased results. One possible example comes from a 2021 study examining COVID-19 mortality among patients admitted to any Veterans Affairs (VA) hospital [21]. While this analysis was not the main objective of the study, a proportional hazard analysis showed that a history of smoking or obesity were both protective against the risk of dying from COVID-19 among this population (see Table 3 of the article) [21]. This correlation is at odds with most of the COVID-19 research carried out during the pandemic, which identifies obesity and a history of smoking as risk factors for COVID-19 related mortality [22-24]. In other words, the findings from this sample of VA patients do not appear to be generalizable. One reasonable hypothesis for this discrepancy in findings is that the characteristics of veterans, and particularly veterans whose illness was severe enough to present to the hospital, differ from those of the broader population [21, 24]. For AI models trained on a very specified population like this, the current Data Cards model would help flag this type of upstream bias by explaining the purpose, origins and methods of data collection.

### Exclusion Criteria

Exclusion criteria are a necessary element of any study design, AI-based or not, but they often limit generalizability [26]. Such criteria can cause selection bias in the original dataset (prior to AI analysis), but AI and machine learning studies themselves may impose additional exclusion criteria. In clinical trials, a systematic review of RCTs found that three-quarters of all trials examined had eligibility criteria that excluded over half of relevant patients, with notable exclusion rates of 83%, 96.0% and 84.3% for hypertension, asthma and COPD, respectively [27]. More generally, frequently utilized exclusion criteria like age, comorbidities, and co-prescribing often ensure that participants in studies are in better health at baseline compared to the broader population [28].

Quantifying the impact of exclusion criteria can be challenging, as outcomes are rarely tracked and/or reported for excluded groups, and exclusion criteria are often not reported [29], particularly in the case of AI studies [30]. Thus, we provide two notable non-AI examples of subsequent analyses of heart disease clinical trial data that show how exclusion criteria can affect a study population. One approach re-analyzed data from the DANAMI-3 multicenter clinical trial investigating ST-elevation myocardial infarction (STEMI) and compared outcomes of participants from comparable registries to those who were not eligible for the trial [31]. They found that patients who were excluded had an approximately 3.42 times greater chance of mortality from a STEMI compared to the trial participants [31]. Similarly, a study comparing 8,469 patients from the Global Registry of Acute Coronary Events registry found that trial participants had lower mortality rates (3.6%) from acute myocardial infarction compared to excluded patients (11.4%) [32]. If either of these datasets were used as part of an AI approach to predict risk of myocardial infarction mortality, the algorithm would likely have an overly optimistic and biased assessment of mortality risk.

### Input-Level Exclusion

One type of exclusion that is more unique to AI studies is exclusion at the input-data level, which can occur due to poor quality or missing data. Data that are missing can be grouped into distinct classifications: data that are Missing Completely At Random (MCAR) has no systematic discrepancies; Missing At Random (MAR) shows systematic absence in observed data but not in unobserved; and Missing Not At Random (MNAR) displays a systematic pattern and can thus be a source of selection bias [33]. In AI studies, there is often either a threshold of missingness allowed, or in some cases, patients with missing data are removed altogether to create curated datasets for training and testing [34]. However, much of the time missing data is simply not addressed, with one study from 2017 finding that 49 out of 107 electronic health record-based risk prediction approaches evaluated did not mention missing data at all [35].

Paying attention to missing rates is critical since they can and often do vary systematically across populations and can thus be classified at MNAR. For example, patients with greater psychosocial needs [36] and patients with low SES [37] are both more likely to seek care at multiple hospitals compared to the average patient. Accordingly, these patients’ data may be spread across multiple health facilities, increasing the likelihood of missing observations in a single-center machine learning dataset. Individuals with low SES and racially minoritized individuals may also tend to use the healthcare system less often due to structural barriers resulting in less access, including receiving less testing, so studies examining an electronic health record would report more missing data for these patients [38].

A recent study using the Medical Information Mart for Intensive Care III (MIMIC-III) database demonstrates that the addition of missing data tends to more negatively impact the performance of disease prediction models for groups that have less access to healthcare [38]. Specifically, when stratified by insurance status, patients insured by Medicare/Medicaid experienced a sharper drop in model performance as the amount of missing data increased compared to those with private insurance [38].

### Patient Attrition (Outcome-Level Exclusion)

Selection bias can also arise even after making final exclusions and inclusions of participants in a study. This commonly happens when patients are lost to follow-up, but also if participants revoke consent or experience a protocol deviation. In AI studies using clinical data, this would manifest as missing outcomes data (as opposed to missing input data noted above).

The vast majority of medical datasets show patient attrition; among manuscripts published in frequently-cited journals in 2014, missing outcomes data was present in nearly 95% of trials, with a median 9% (range of 0-70%) of patients per trial with a missing outcome [39]. To address this issue, researchers often employ techniques such as time-to-event modeling with censorship or utilize sensitivity analyses, including the fragility index which gauges the minimal number of patient results that, if changed, would reverse a study’s statistical significance [40]. Intriguingly, an application of this index found that in 53% of trials, the fragility index was fewer than the number of participants lost to follow-up with some studies having fragility indices of only 1 participant [40].

Despite their utility in evaluating robustness of study findings, these approaches cannot capture how disproportionate participant dropout by clinical or sociodemographic characteristics may affect either study results or the subsequent bias of AI algorithms that filter out this missing outcomes data. This is an issue given that in some clinical and research contexts, rate of attrition may differ by demographic and clinical features. For example, non-white race, male sex, and lower functional status were all found to be significantly associated with a higher probability of loss to follow-up after hip arthroscopy [41]. Furthermore, a review of HIV trials identified lower retention rates in Black patients [42] while another approach found an association between speaking a language other than English and increased likelihood of dropout in a cervical cancer clinical trial [43]. Ultimately, these examples suggest that missing follow-up data is often MNAR, which can lead to systematic bias in AI-based algorithms training on this data.

## Recommendations for AI Researchers

We have discussed how recruitment, exclusion criteria, input- and outcome-level exclusions can all lead to selection bias, resulting in a study sample that is discordant from the target population. This selection bias can then be encoded by an algorithm that trains on that clinical dataset, leading to models that perform well on the types of patients it has seen but less so on those it has not, leading to inequity. Given that medical AI approaches often utilize cohorts from RCTs and cohort studies and perform their own processing steps before using them as training, validation and testing datasets, it is crucial to standardize the reporting of these steps to inform readers and users of possible biases.

Before performing any data processing or modeling, we first recommend that AI researchers assess their training and testing datasets and consider the ways in which they may be under-counting certain populations including through previous application of exclusion criteria to the dataset. The Data Card initiative can be a major part of this assessment, as it necessitates the reporting of dataset features, including its origins, the populations represented, and its prior processing [13]. Furthermore, we strongly recommend that researchers think critically about what sociodemographic and clinical factors are relevant to be tracked for their study, balancing knowledge of structural disparities in health with acknowledgement that characteristics like race and gender are socially constructed. Rather than serving as passive reporters, researchers should seek to identify why disparities in study exclusion, treatment access, and follow up occur, and how they can be prevented in the future.

The subsequent step involves monitoring how the composition of a cohort changes throughout an AI study itself to prepare the data for modeling. We introduce an updated participant flow diagram tailored for AI research that we hope to serve as a standardized best practice method for tracking sample composition – see Figure 1. This approach not only complements the Data Card initiative but also has the potential to be a benchmark reporting technique alongside the traditional “Table One” used in clinical studies to outline the final and post-processed data characteristics. Specifically, we visualize cohort composition and shifts in demographic details such as age, gender, socioeconomic status and race or ethnicity. Depending on the study design, researchers could also choose to track selected clinical characteristics through this diagram, such as the SOFA score in a critical care study, to ensure they are not altered significantly.

**Figure 1:**
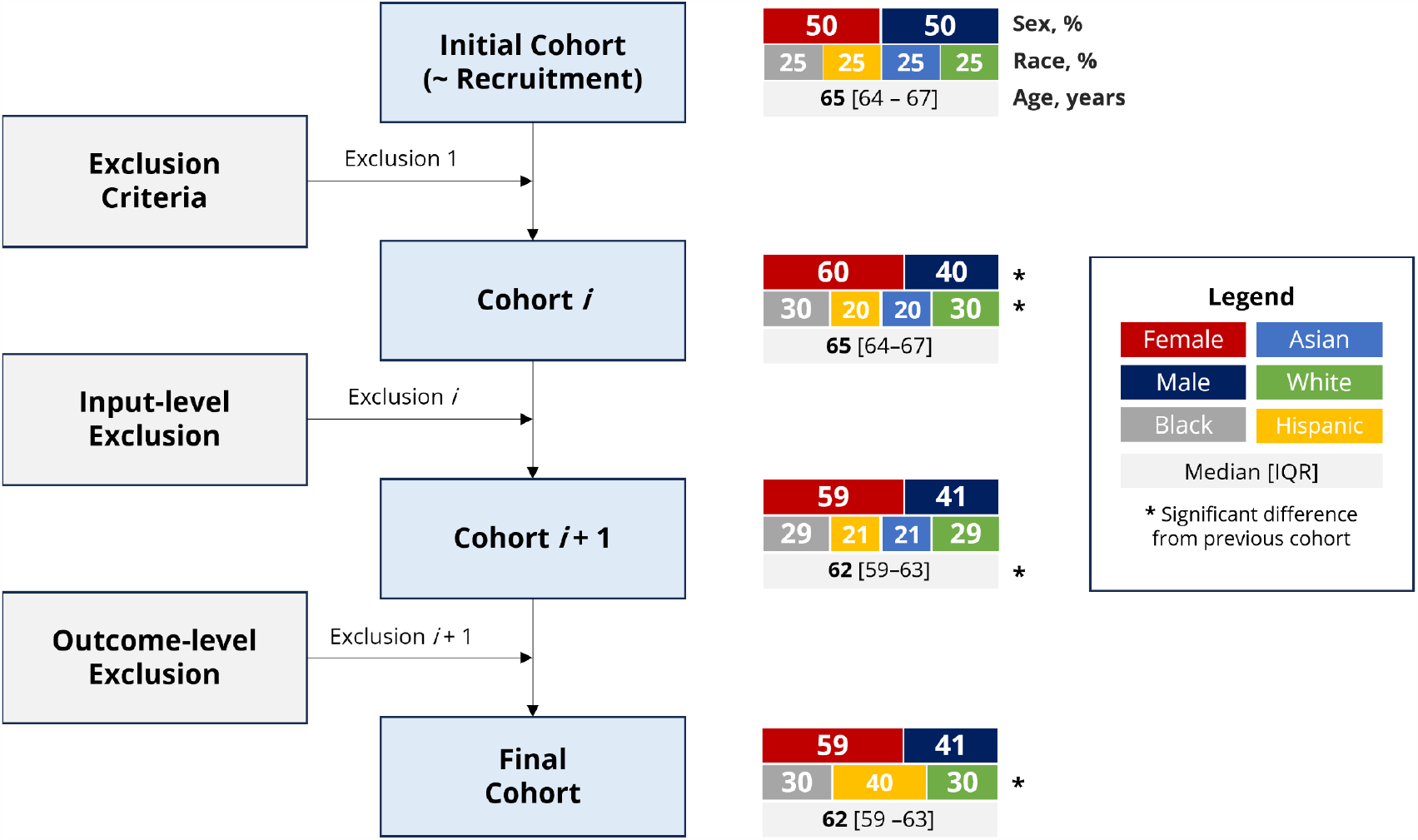
Template of the updated Participant Flow Diagram.

We further include the reporting of standardized mean difference (SMD) as a statistical method of measuring effect size in the diagram to ensure that the demographics of the excluded patient population do not differ in a clinically significant manner from the study sample after a given step [44]. Borrowing from literature on inverse probability of treatment weighting, we suggest that a SMD greater than an absolute value of 0.1 between steps may represent a meaningful change in sample composition [45]. Alternative methods of comparison such as significance testing may be considered by investigators, however care should be taken to consider type II error in large analytic samples and type I error in small samples when comparing changes in sample composition using p-values.

This standardized depiction not only showcases the evolution of a sample throughout a study but also promotes the reporting of clinically-relevant characteristics that may serve as sources of confounding. The routine use of this diagram can help to uncover many currently hidden biases that occur as a study progresses and help to ensure that the results of a clinical algorithm are not improperly extrapolated to excluded populations.

## Example Case Study

Pulse oximetry is a prominent example of how racial and ethnic bias can manifest in critical care medical equipment [46]. Underperformance of the pulse oximeter in patients with darker skin color has been shown to result in events of hidden hypoxemia, which can be defined as SaO_2_ (measured by arterial blood gas [ABG]) < 88%, but SpO_2_ (measured by pulse oximetry) ≥ 92% [45, 46].

We provide a hypothetical case study here of the development of an ML model to predict the likelihood of the presence of hidden hypoxemia based on a patient’s oxygen saturation trajectory since ICU admission. Specifically, we discuss the sample selection process for training such a model using real data from the MIMIC-IV dataset [49]. However, the creation of such models can be limited by a disproportionate absence of ABG measurements, resulting in a higher rate of exclusion of Black patients [50].

We focus on each individual step of the sample selection process below before illustrating how the updated patient flow diagram can illuminate some of the issues that arise during this process.

1. **Recruitment:** The database selection serves as the Recruitment phase here. The first step is to examine MIMIC-IV, a single-center Electronic Health Records (EHR) database from Boston, captured between 2008 and 2019 [49]. This dataset comprises ICU patients – a population that was able to reach the hospital to receive critical care in the first place – introducing a potential selection bias that should be noted in any study using this dataset. Algorithms that are trained on MIMIC-IV data should target a population that is adequately represented in the dataset, otherwise both dataset and subpopulation shifts may occur [49, 50].
2. **Exclusion Criteria:** An SpO_2_ ≤ 92% should trigger supplemental oxygen administration in the ICU. To have a model focused on patients at harm of developing hidden hypoxemia, we will limit our cohort to patients with SpO_2_ > 92%.
3. **Input-level exclusion:** Patients with missing SpO_2_ values, outliers (e.g., out-of-range oxygen saturation), or without relevant demographic information should be removed from the study.
4. **Outcome-level exclusion:** Patients without an ABG that can serve as the ground-truth for this ML algorithm predicting future hidden hypoxemia should also be removed. The presence of hidden hypoxemia is assessed when comparing SaO_2_ and SpO_2_ paired within 5 minutes; patients will be excluded if the pairing is not possible.

The participant flow diagram describing these steps and their effects on the cohort composition is depicted in Figure 2. Notably, the order of applying the aforementioned exclusion criteria is not fixed, and depends on the data structure. In this case, for implementation efficiency, the criteria were applied in reverse order, starting with the outcome-level exclusion.

**Figure 2:**
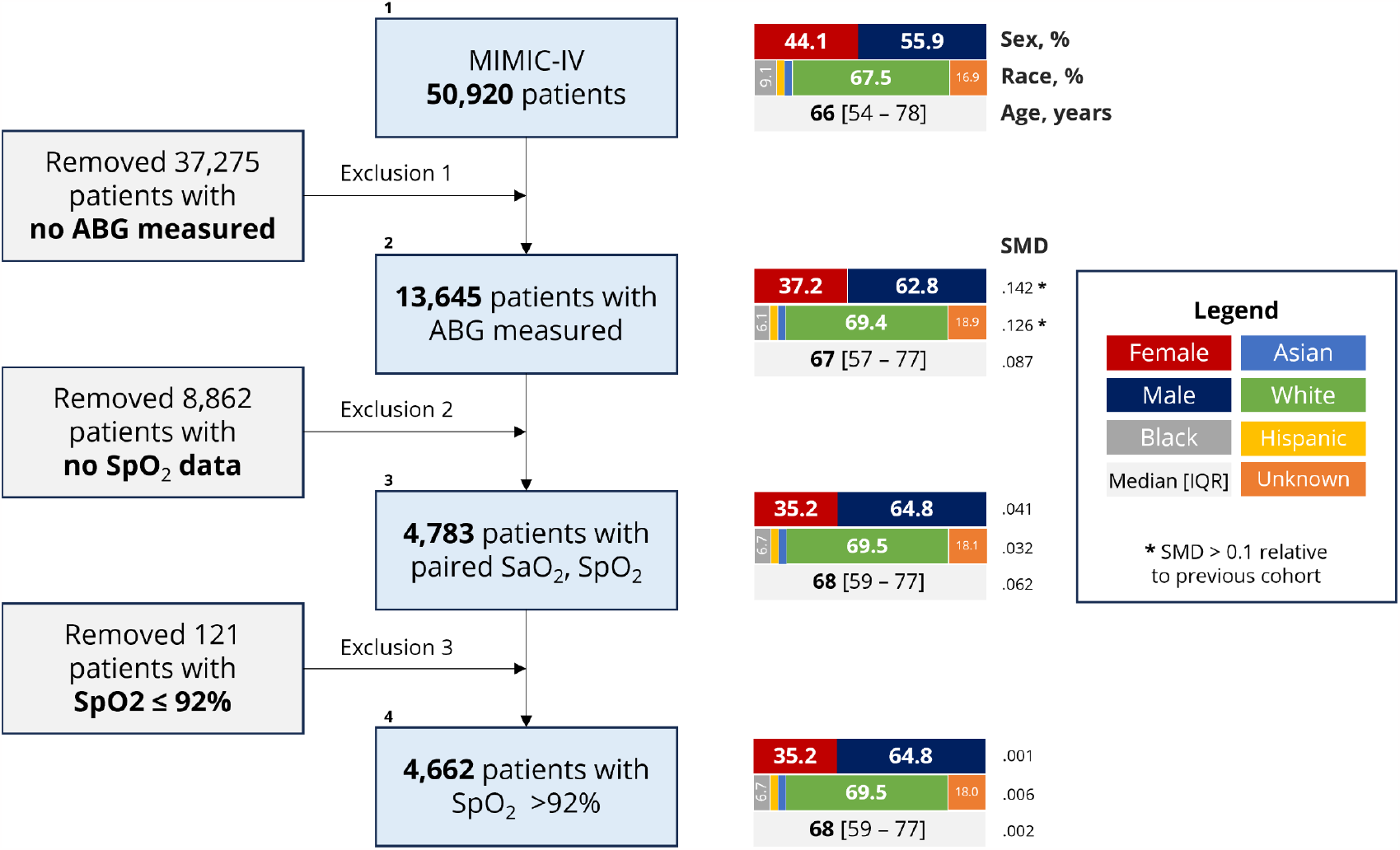
Example Case Study’s Participant Flow Diagram.

In the outcomes-level exclusion of this study (removing patients without an ABG), there is a clear shift in the cohort composition with a change of 55.9% to 62.8% in the percentage of males as well a shift from 67.5% to 69.4% White patients (Figure 2; full data summary in Table 1). There is also a notable drop in Black patients from 9.1% to 6.1% after this step (Table 1). The above shifts were flagged by our diagram, and can be shown by the measured SMD for these two categories. These changes could result for a number of reasons, which include certain demographics being less likely to receive an ABG measurement in the hospital [50]. The participant flow diagram allows us to see the disproportionate dropout and to inform the study and the findings, which could lead to an investigator adjusting the study as a whole to lead to more inclusion or reporting this as a key limitation depending on the situation. Finally, this method of tracking also has the potential to illuminate disparities and differences between populations we may not have not known existed prior.

**Table 1.** Auxiliary Table Showing Case Study Results.

|  |  | Cohort Number (Step of the Flow Diagram) |  |  |  | Effect Size |  |  |
| --- | --- | --- | --- | --- | --- | --- | --- | --- |
|  |  | 1 | 2 | 3 | 4 | SMD (1,2) | SMD (2,3) | SMD (3,4) |
| **n** |  | 50,920 | 13,645 | 4,783 | 4,662 |  |  |  |
| **Race and Ethnicity, n (%)** | **Asian** | 1,496 (2.9) | 332 (2.4) | 112 (2.3) | 112 (2.4) | .126 | .032 | .006 |
|  | **Black** | 4,640 (9.1) | 838 (6.1) | 320 (6.7) | 311 (6.7) |  |  |  |
|  | **Hispanic/Latino** | 1,783 (3.5) | 424 (3.1) | 161 (3.4) | 159 (3.4) |  |  |  |
|  | **White** | 34,382 (67.5) | 9,472 (69.4) | 3,324 (69.5) | 3,242 (69.5) |  |  |  |
|  | **Unknown** | 8,619 (16.9) | 2579 (18.9) | 866 (18.1) | 838 (18.0) |  |  |  |
| **Sex, n (%)** | **Female** | 22,480 (44.1) | 5,075 (37.2) | 1,684 (35.2) | 1,639 (35.2) | .142 | .041 | .001 |
| **Age, median [Q1,Q3]** | **Admission** | 66.0 [54,78] | 67.0 [57,77] | 68 [59,77] | 68 [59,77] | .087 | .062 | .002 |

## Conclusion

Our proposed approach and updated participant flow diagram aims to identify possible biases in AI data processing, broaden the understanding of a study’s applicability, and promote researcher accountability in eliminating structural biases in biomedical research. We intend this to be a useful tool and overview of ways that bias can enter studies at any stage, but we acknowledge this approach will not by itself remove bias from studies. Further, a key limitation is that this approach will not address algorithmic encoding of structural biases already present in a dataset, such as in the case of physician implicit bias [53]. Necessary future work includes the development of an easily implementable tool that can generate this diagram automatically and in a standardized manner for any study. Finally, while we discuss many of the implications for AI and machine learning studies here, these same lessons of tracking excluded participants apply equally to clinical RCTs and observational studies.

## Data Availability

All data used are available online at https://physionet.org/content/mimiciv/2.2

## Notes

**Data Availability** The data that support the example case study are available in MIMIC-IV with the identifier doi.org/10.1093/jamia/ocx084 publicly available on PhysioNet (https://physionet.org/content/mimiciv/2.2/).

**Conflicts of Interest** All authors report no conflicts of interest.

### Competing Interest Statement

The authors have declared no competing interest.

### Funding Statement

This research did not receive any specific grant from funding agencies in the public, commercial, or not-for-profit sectors.

### Author Declarations

The study used ONLY openly available human data that were originally located at PhysioNet.

